# Analysis of the Association between Sarcopenia Parameters and Falls in Patients with Chronic Kidney Disease undergoing Hemodialysis: A Cross-Sectional Study

**DOI:** 10.1101/2025.07.01.25330659

**Authors:** João Vitor Oblanca, Thaís Larissa Reichert, César Faúndez-Casanova, Kauana Borges Marchini, Fábio Torres, Mariana Ardengue, Sergio Seiji Yamada, Ademar Avelar

## Abstract

**Introduction:** Sarcopenia has been associated with an increased risk of falls in diverse populations. Patients with end-stage kidney disease (ESRD) undergoing hemodialysis (HD) have an increased prevalence of muscle weakness and wasting. The aim of this study was to investigate the association between parameters of sarcopenia and a history of falls in ESRD patients in HD.

**Methods:** A cross-sectional study design was utilized to assess 111 participants with ESRD in HD (54 ± 15.6 years; 59.5% men) in a tertiary hospital in Maringa-PR. Sarcopenia status was determined through the measure of muscle strength and mass. Muscle strength was assessed through hand grip strength utilizing a handheld dynamometer. Muscle mass was estimated through bioelectrical impedance. Falls’ history was assessed with the fifth question of the SARC-F questionnaire. Data normality was assessed with the Shapiro-Wilk test. The association of the sarcopenia parameters with a falls history was analyzed with the chi-square test and Cramér’s *V* to measure effect size.

**Results:** No association was found among the parameters of sarcopenia or its diagnosis and a history of falls in this population (*p* > 0.05). Participants who reported falling in the previous year were older (60 ± 16.7 *vs.* 51 ± 14.7 years, *p* = 0.01) and had a history of cerebrovascular disease (14.3% *vs.* 2.4%, *p* = 0.02).

**Conclusion:** Falls history was not associated with muscle strength or mass in ESRD patients in HD. Future longitudinal studies are needed to investigate other factors associated with this outcome.

## Introduction

Around 850 million people worldwide live with some form of chronic kidney disease (CKD) which is characterized by the presence of one or more markers of kidney damage and a decreased glomerular filtration rate (GFR) (<60 ml/min/1.73 m^2^) present for a minimum of three months^1^. CKD is categorized by the GFR and those who present with values <15 have end stage renal disease (ESRD) and necessitate renal replacement therapy (RRT) as part of their ongoing treatment plan^1^. Most patients with ESRD in Brazil undergo hemodialysis (HD) as their RRT of choice, with over 150 thousand people in treatment in 2022^2^. CKD can be associated with complications such as hypertension, cardiovascular disease, bone mineral disorder, metabolic acidosis, and uremic symptoms, among others^3^. Common complications associated with HD treatment are intradialytic hypotension and muscle cramps, which can predispose a patient to experiencing a fall^4^.

Falls occur when a person unintentionally comes to rest on the floor, the ground or any other lower level^5^. In a study with Brazilian HD patients it was found that 37.4% had fallen at least once in the previous year^6^. Older CKD patients fall more often than younger patients^7^. Falls increase the risk of hip and nonvertebral fractures in HD patients^8^. The use of walking aids and the presence of cardiovascular or cerebrovascular conditions increase the risk of falling in HD patients^9^. The post-dialysis period, the number and type of medications used, increased postural sway, and fear of falling are known risk factors for falling in this population^10–15^. Diagnostic parameters of sarcopenia, such as low muscle strength and decreased physical function are also associated with falls in ESRD patients in HD^16^.

Among the factors that increase the risk of falls, sarcopenia, a generalized and progressive skeletal muscle disorder which is characterized by the accelerated loss of muscle mass and function, stands out^17^. The prevalence of sarcopenia in the general population is 5% to 10% while studies in the HD population estimate that between 9.8% and 28.5% of patients present with the disorder, depending on the criteria used for the definition^18–20^. A diagnosis of sarcopenia is associated with an increased risk of mortality and cardiovascular (CV) events in CKD patients and in those undergoing HD^21–23^. Slow gait speed and low hand grip strength are independent predictors of fatal and non-fatal CV events in HD patients^24,25^.

Besides increasing the predisposition to falls, sarcopenia increases their overall risk of mortality, especially in community-dwelling older people^26–28^. The physiopathological changes associated with ESRD and HD treatment predispose this population to falls, though the studies conducted have had small sample sizes and included mostly elderly patients^14,16,31^. Due to the increased risk of mortality and the prediction of an increase in the prevalence of CKD patients for the following years, the impact of sarcopenia and falls in HD patients warrants further study^1^.

The screening of HD patients by a multidisciplinary team for signs of sarcopenia is important for successful care^29^. Assessing physical function parameters such as gait speed and step length, identifying patients with a fear of falling and creating safe environments for care help to minimize fall risk^30–32^.

Taking these steps can lead to an increased quality of life and the maintenance of functional independence during treatment^33^. Therefore, the aim of this study was to investigate the association between low muscle strength, low muscle mass and sarcopenia diagnosis with falls in CKD patients in HD. The hypothesis is that those participants who present with low muscle strength, low muscle mass, and a diagnosis of sarcopenia would report more falls in the previous year.

## Methods

### Study Design

This was a single-center, cross-sectional, and observational study. The manuscript was prepared following the STROBE (Strengthening the Reporting of Observational Studies in Epidemiology) statement to ensure comprehensive and transparent reporting^34^. The study was approved by the research ethics committee of the Universidade Estadual de Maringa under protocol number 6.004.620 and was conducted in accordance with the Declaration of Helsinki. The project was also submitted for review and approval by the ethics committee of the hospital responsible for the HD center, with authorization granted for the use of clinical facilities and access to the research participants’ data. All study participants provided written informed consent before enrollment.

### Setting

The study was conducted at the nephrology unit of Hospital Santa Casa de Maringa (HSCM) located in Maringá, state of Paraná, Brazil. Participant recruitment and data collection took place between May and December of 2023.

### Participants

The population of this study was composed of patients with ESRD undergoing HD who received care at the nephrology unit of HSCM. The participants of this study are also part of the larger project “Assessment and monitoring of global health and survival of people with chronic kidney disease undergoing hemodialysis: a cohort study” which was approved in Public Call No. 421091/2023 - CNPq. A non-probabilistic, convenience sample was utilized in this study. In order to reduce potential sources of bias, such as response bias from self-reported data, all research staff were previously trained on how to conduct questionnaires and physical exams.

The inclusion criteria were: ≥ 18 years of age; diagnosis of chronic kidney failure; ≥1 month of HD treatment at HSCM; able to comprehend the questionnaire and participate in the physical exam; being clinically stable (absence of hemodynamic instability, need for intensive care and/or any acute decompensated condition). The exclusion criteria were: being permanently bedridden without the ability to walk; placed in isolated treatment due to active infection; and having any limb amputation.

### Variables

#### Sociodemographic and health data

Data were collected from all participants included in the study by means of a standardized questionnaire. The data collected were: age, gender, race, marital status, highest level of education achieved, household income, alcohol consumption, and smoking status. Patients height (m), weight (kg), HD vintage and other comorbidities were collected from their medical records. Body mass index (BMI) was calculated by dividing weight by height squared (kg/m^2^).

#### Sarcopenia

The assessment and confirmation of sarcopenia status was realized with the definition in the updated version of the European Working Group on Sarcopenia in Older People (EWGSOP2)^35^. The assessment of sarcopenia status is obtained through the measurement of muscle strength and confirmation is achieved by assessing muscle quantity or quality.

#### Muscle strength

Muscle strength was assessed through a handgrip strength (HGS) test utilizing a hand held digital dynamometer (Saehan SH001). Participants were tested in a seated position, with both feet on the ground, shoulder slightly abducted, elbow flexed to 90°, and forearm in a neutral position. HGS was assessed three times alternatively in both arms with one minute of rest between each set. Participants were instructed to squeeze the dynamometer with their maximum strength and hold for five seconds with the highest value obtained recorded. The low HGS cut-off values determined by the EWGSOP2 are <27 kg and <16 kg for men and women, respectively^35^. The participants who failed to reach these threshold values with their right hand were classified as having low muscle strength.

The chair stand test (CST) was used to assess lower limb muscle strength. Participants began the test sitting down in a chair with their arms crossed on their chest, keeping their feet flat on the floor, and their backs straight. They were instructed to rise to a full standing position, then sit back down again five times. The time taken to complete the test was measured with a digital chronometer. Participants repeated the test twice and the fastest time taken to complete it was recorded. As per the EWGSOP2 cut-off points for the chair stand test, those who took >15 seconds to complete the test were classified as having low muscle strength^35^. Participants who presented with low muscle strength in either test were classified as having probable sarcopenia as per the EWGSOP2.

#### Muscle mass

Lean mass was estimated through bioelectrical impedance analysis (BIA). Participants were instructed to refrain from extraneous physical activity and to avoid consuming alcoholic and/or caffeinated beverages 24 hours before the assessment. Participants were evaluated with standard equipment (BIA Analyzer^™^) in a supine position and electrodes were placed five centimeters apart on their right hand and right foot. The resistance (Rz), reactance (Xc) and phase angle values obtained were recorded and were utilized with the following formula in order to determine the participants appendicular lean mass (ALM)^36^:

*ALM = −3.964 + (0.227 x Ri) + (0.095 x weight (kg)) + (1.384 x sex) + (0.064 x XC)*
where RI is the resistive index (height in centimeters squared/Rz) and sex is 1 for men and 0 for women. The participants’ ALM was further corrected by their height squared (m^2^). The cut-off values for low ALM/m^2^ are <7.0 kg/m^2^ for men and <5.5 kg/m^2^ for women, as determined by the EWGSOP2 criteria^35^. Sarcopenia diagnosis was confirmed for those participants who presented with both low muscle strength and low muscle quantity and participants were divided into two groups: non-sarcopenic and sarcopenic.

#### SARC-F

The SARC-F is a five item questionnaire that is used as a screen for sarcopenia risk. Participants were asked about their perceived limitations in strength, walking ability, rising from a chair, climbing stairs and experiences with falls in the past year^35^. The fifth question (‘How many times have you fallen in the past year?’) has three possible answers: a) none; b) one to three falls; and c) four or more falls. Patients were divided into two groups based on their answer: non-fallers and fallers.

### Procedures

All participant data were collected by trained and experienced professionals. In order to minimize the possible effects of hemodynamic instability, participants were only assessed on the second and third day of their scheduled HD sessions. Muscle strength and mass assessments were conducted prior to the HD session while sociodemographic and health data were collected during the session.

### Statistical Analysis

Statistical analyses were conducted using JASP version 0.19.3. Descriptive statistics were used to summarize the sample characteristics. Continuous and categorical variables are presented as mean ± standard deviation (SD), median and 25^th^ and 75^th^ percentiles, and percentages, respectively. Data normality was assessed with the Shapiro-Wilk test and homogeneity was assessed with Levene’s test. The comparison between the groups of non-fallers and fallers were realized with Student’s *t* test, Welch’s *t* test or Mann-Whitney *U* test for independent variables according to normality and homogeneity. Cohen’s *d* was used to estimate the effect sizes (small = 0.2, medium = 0.5, and large ≥ 0.8). The associations between categorical variables was assessed with the χ^2^ test and the effect size was estimated using Cramér’s *V* (1 df_min_ = small (0.1), moderate (0.3), and large (0.5))^37^. Missing data were handled using complete case analysis. A two-sided *p*-value <0.05 was considered statistically significant. Any missing data were eliminated from the analysis.

## Results

There were 244 patients in the HD unit of HSCM who were potentially eligible for inclusion in the study, of which 77 did not meet the inclusion criteria for the study. The remaining 167 participants consented to participate in the study and their data was collected. The data from 56 of these participants was not included in the final analysis due to missing variables (missing CST data = 50; missing ALM data = 14; missing HGS data = 7; missing SARC-F data = 4; and missing body mass data = 1). Some participants had more than one variable missing, and thus the resulting missing data number exceeds the number of participants excluded. The final sample was made up of data from 111 participants (Figure 1).

**Figure 1.**
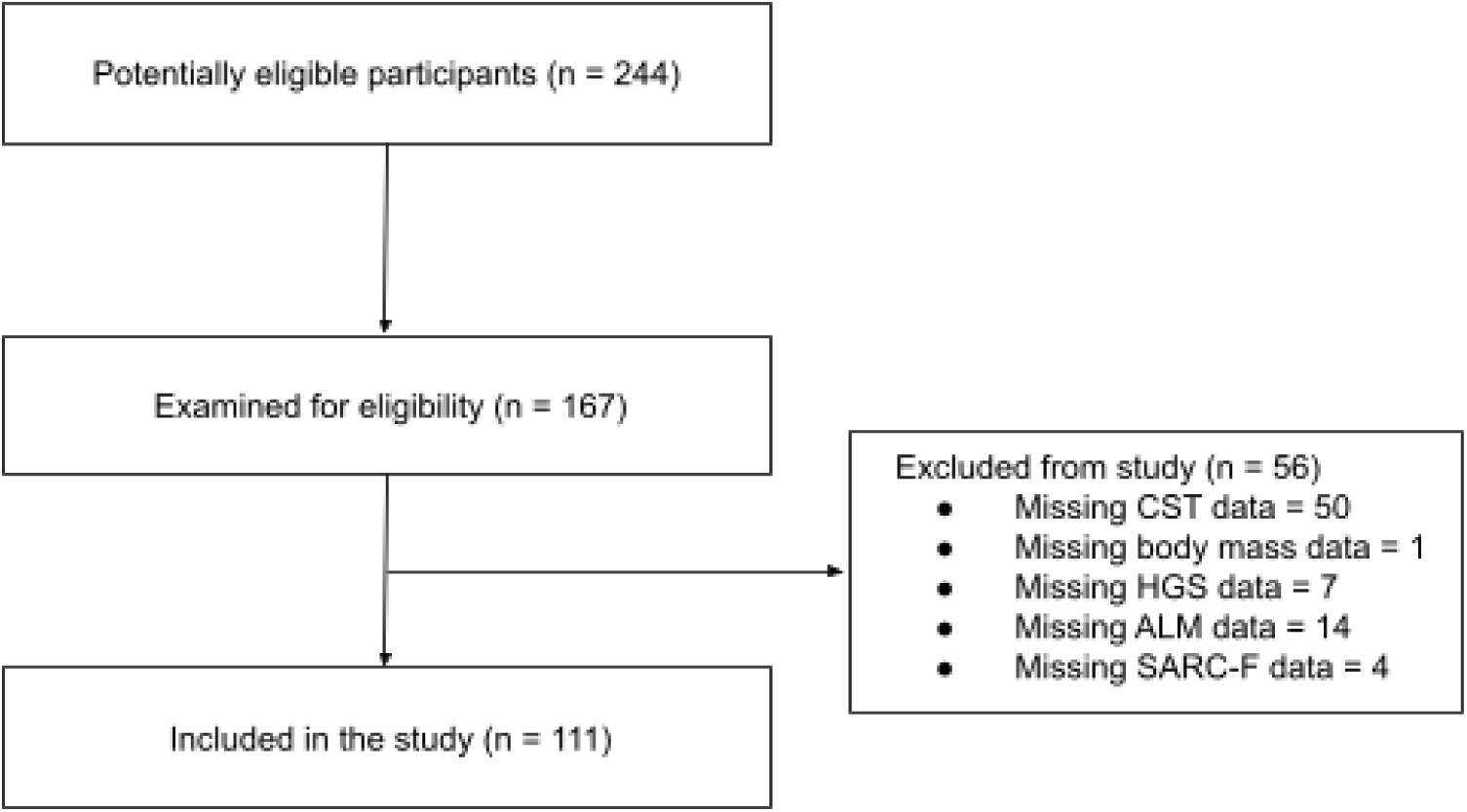
STROBE flow chart.

Participants’ sociodemographic and clinical characteristics are described in Table 1. Falls in the previous year were reported by 25.2% of the sample (28/111). These participants were grouped together in the fallers group, while the remaining were classified as non-fallers. Fallers were significantly older (60 ± 16.7 years *vs*. 51 ± 14.7 years, *t* = −2.67, *p* = 0.01) than non-fallers. There was a mean difference of −8.9 years with a 95% confidence interval of −15.5 to −2.3 years between the groups, with a medium to large effect size (Cohen’s *d* = 0.58). Those with a previous clinical history of stroke were also 1.9 times more likely to report a fall in the previous year (14.3% *vs*. 2.4%, χ^2^ = 5.78, *p* = 0.02, OR = 1.91 (0.15 - 3.67). Those who reported a fall showed a trend for having less ALM/m^2^ (6.6 kg/m^2^ (6.3 - 7.1) *vs*. 7.1 kg/m^2^ (6.3 - 7.7)) but it did not reach statistical significance (*p* = 0.052). 46.4% of participants who experienced a fall had probable sarcopenia, while 17.9% had confirmed sarcopenia status; these values were not significantly different from participants who did not fall. There were no significant differences between the groups for any other sociodemographic or clinical characteristic.

**Table 1.** Sociodemographic and Clinical Characteristics of the Sample.

|  | Total (n = 111) | No Falls (n = 83) | Falls (n = 28) | *p* |
| --- | --- | --- | --- | --- |
| Age (years) (M ± SD) | 54 ± 15.6 | 51 ± 14.7 | 60 ± 16.7 | 0.01* |
| Gender (%) |  |  |  |  |
| *Male* | 59.5 | 57.8 | 64.3 | 0.547 |
| *Female* | 40.5 | 42.2 | 35.7 |  |
| Height (m) (M ± SD) | 1.66 ± 0.09 | 1.66 ± 0.09 | 1.66 ± 0.09 | 0.85 |
| Body mass (kg) (M ± SD) | 71.5 ± 15.6 | 72.7 ± 16.8 | 68.2 ± 11.1 | 0.12 |
| BMI (kg/m<sup>2</sup>) (Median (P25 - P75)) | 24.6 (22.5 - 28.2) | 25.5 (22.5 - 28.7) | 23.8 (22.4 - 25.8) | 0.19 |
| HD vintage (months) (Median (P25 - P75)) | 30.0 (12.0 - 57.5) | 32.0 (16.0 - 60.0) | 19.5 (10.8 - 50.5) | 0.19 |
| HGS (kg) (Median (P25 - P75)) | 26.9 (19.7 - 35.6) | 26.9 (20.8 - 36.6) | 26.4 (18.5 - 35.1) | 0.57 |
| CST (sec) (Median (P25 - P75)) | 11.8 (9.4 - 15.2) | 11.7 (9.3 - 15.0) | 12.6 (11.0 - 15.8) | 0.34 |
| ALM/m<sup>2</sup> (kg/m<sup>2</sup>) (Median (P25 - P75)) | 7.0 (6.3 - 7.6) | 7.1 (6.3 - 7.7) | 6.6 (6.3 - 7.1) | 0.05 |
| Marital status (%) |  |  |  |  |
| *Single* | 19.8 | 21.7 | 14.3 | 0.30 |
| *Married* | 56.8 | 55.4 | 60.7 |  |
| *Cohabiting* | 6.3 | 8.4 | 0.0 |  |
| *Divorced* | 13.5 | 10.8 | 21.4 |  |
| *Widowed* | 2.7 | 2.4 | 3.6 |  |
| Education level (%) |  |  |  |  |
| *Incomplete primary education* | 39.6 | 36.1 | 50.0 | 0.55 |
| *Complete primary education* | 13.5 | 14.5 | 10.7 |  |
| *High school diploma* | 28.8 | 31.3 | 21.4 |  |
| *Bachelor's degree* | 12.6 | 12.0 | 14.3 |  |
| *Postgraduate diploma* | 3.6 | 4.8 | 0.0 |  |
| *Master's degree* | 1.8 | 1.2 | 3.6 |  |
| Household income (%) |  |  |  |  |
| *Up to 1 minimum wage* | 15.3 | 15.7 | 14.3 | 0.13 |
| *From 1 to 2 minimum wages* | 28.8 | 25.3 | 39.3 |  |
| *From 2 to 6 minimum wages* | 34.2 | 39.8 | 17.9 |  |
| *From 6 to 10 minimum wages* | 10.8 | 8.4 | 17.9 |  |
| *More than 10 minimum wages* | 8.1 | 9.6 | 3.6 |  |
| Alcohol use (%) | 18.9 | 20.5 | 14.3 | 0.47 |
| Tobacco use (%) | 10.8 | 9.6 | 14.3 | 0.49 |
| Hypertension (%) | 90.1 | 90.4 | 89.3 | 0.87 |
| Diabetes (%) | 34.2 | 31.3 | 42.9 | 0.27 |
| Dyslipidemia (%) | 27.0 | 28.9 | 21.4 | 0.44 |
| Heart failure (%) | 9.9 | 7.2 | 17.9 | 0.10 |
| Cancer (%) | 2.7 | 2.4 | 3.6 | 0.74 |
| Stroke (%) | 5.4 | 2.4 | 14.3 | 0.02* |
| Probable Sarcopenia (%) | 43.2 | 42.2 | 46.4 | 0.69 |
| Confirmed Sarcopenia (%) | 12.6 | 10.8 | 17.9 | 0.33 |

Note: M = mean; SD = standard deviation; BMI = body mass index; P25 = 25<sup>th</sup> percentile; P75 = 75<sup>th</sup> percentile; HD = hemodialysis; HGS = hand grip strength; CST = chair stand test; ALM/m<sup>2</sup> = appendicular lean mass divided by height squared. Student's *t* test used for age and height; Welch's *t* test used for body mass; Mann-Whitney *U* test used for BMI, HD vintage, HGS, CST, and ALM/m<sup>2</sup>; Chi-squared test used for gender, marital status, education level, household income, alcohol use, tobacco use, hypertension, diabetes, dyslipidemia, heart failure, cancer, and stroke, probable sarcopenia, and confirmed sarcopenia. \**p* < 0.05.

The associations between the presence of falls in the past year and the parameters of sarcopenia are shown in Table 2. A positive association was found between falls reported in the past year and muscle mass status (χ^2^ = 1.13) though it did not reach statistical significance (*p* = 0.29). This association had a small effect size (*V* = 0.10). There were no other significant associations found between the presence of falls and other parameters of sarcopenia.

**Table 2.**
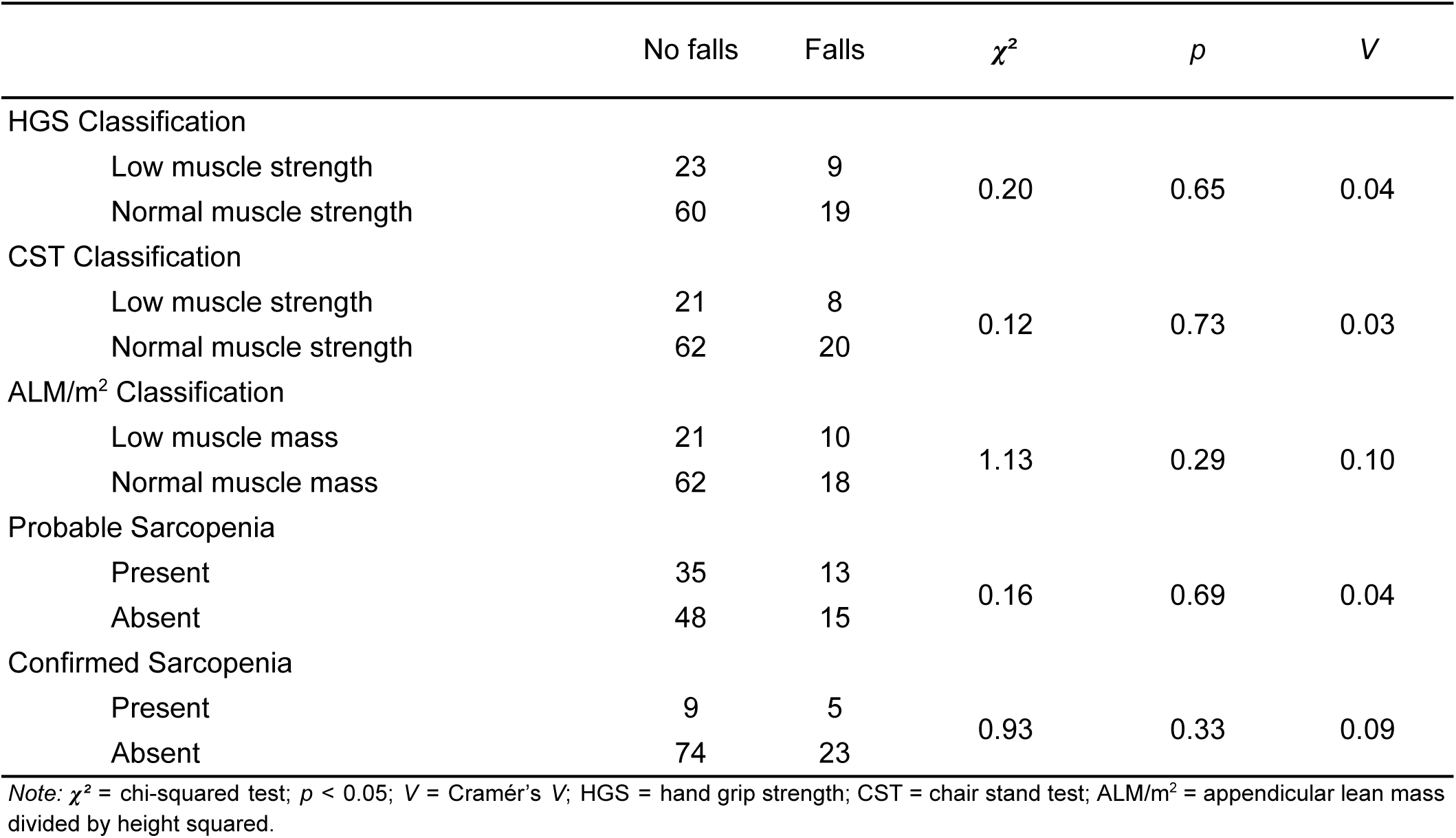
Association Between the Presence of Falls in the past year and Parameters of Sarcopenia.

| | No falls | Falls | $\chi^2$ | $p$ | $V$ |
| --- | --- | --- | --- | --- | --- |
| HGS Classification |  |  |  |  |  |
| Low muscle strength | 23 | 9 | 0.20 | 0.65 | 0.04 |
| Normal muscle strength | 60 | 19 |  |  |  |
| CST Classification |  |  |  |  |  |
| Low muscle strength | 21 | 8 | 0.12 | 0.73 | 0.03 |
| Normal muscle strength | 62 | 20 |  |  |  |
| ALM/m<sup>2</sup> Classification |  |  |  |  |  |
| Low muscle mass | 21 | 10 | 1.13 | 0.29 | 0.10 |
| Normal muscle mass | 62 | 18 |  |  |  |
| Probable Sarcopenia |  |  |  |  |  |
| Present | 35 | 13 | 0.16 | 0.69 | 0.04 |
| Absent | 48 | 15 |  |  |  |
| Confirmed Sarcopenia |  |  |  |  |  |
| Present | 9 | 5 | 0.93 | 0.33 | 0.09 |
| Absent | 74 | 23 |  |  |  |

Note: $\chi^2$ = chi-squared test; $p < 0.05$ ; $V$ = Cramér's $V$ ; HGS = hand grip strength; CST = chair stand test; ALM/m<sup>2</sup> = appendicular lean mass divided by height squared.

## Discussion

This study aimed to investigate the association between low muscle strength, low muscle mass and sarcopenia diagnosis with falls in CKD patients in HD. We found that 25.2% of participants reported falling at least one time in the previous year. These participants who fell were also found to be significantly older and more likely to have a previous medical history of stroke than those who did not report falls. There was a small association found between muscle mass status and a history of falls, though it was not statistically significant.

The prevalence of falls found in our study (25.2%) is similar to that found by Ishii and colleagues^9^ (21.2%) whose study explored the factors related to falls that occurred in older ESRD patients in the HD unit (M = 68.7 years). The authors reported the number of falls in the previous year listed in 629 patients’ medical records. Furthermore, they reported that fallers had a statistically higher chance of having a history of stroke (29.3% in fallers *vs*. 15.5% of non-fallers, *p* = 0.001), similar to what was found in the present study (14.3% in fallers *vs*. 2.4% of non-fallers, *p* = 0.02). Other studies reported a higher prevalence than was found in our study.

Carvalho and Dini^6^ reported that 37.4% of 131 ESRD patients in HD had experienced at least one fall in the previous year. The patients included in their study were similar in age to those found in our study (M = 56.1 years) and they reported fewer cases of hypertension among their cohort (66.3% in non-fallers and 33.6% in fallers) compared to ours (90.4% in non-fallers and 89.3% in fallers). Zanotto and colleagues^38^ found that 37.7% of 69 participants included in their cohort prospectively reported falling at least once during 12 months. Participants included in their study were older than those of our study (M = 61.7 years), while surprisingly they reported that the participants who fell were younger (M = 58.3 years).

Shirai and colleagues^16^ also reported a higher prevalence of falls among those included in their cohort (47.7% of 65 participants). Those with a history of falls were also found to be older (Non-fallers = 71.0 years *vs*. Fallers = 76.0 years, *p* = 0.55), though unlike our study, they did not find a significant difference between the groups. The authors also compared the muscle mass, strength, and physical function data and its impact on the frequency of falls between the groups. Similarly to what was found in our study, muscle mass was not found to significantly impact this outcome.

Unlike the results of this study, Matsumoto and colleagues^39^ found that after adjusting for multiple risk factors the hazard ratio of sarcopenia was more than five times higher in those who reported falling in the previous year. This result suggests that utilizing the fifth question of the SARC-F questionnaire may not be the most appropriate manner to estimate the risk of falls in populations with chronic conditions.

This study had a couple of strengths. First, the use of validated measures of strength and muscle mass with the appropriate cut off values ensured the proper estimation of these variables. Second, the large sample permitted the correct estimation of statistical significance of the associations of sarcopenia parameters and falls. Finally, the use of the tools and validated questionnaires ensured real world applicability due to their affordability and wide availability. This study also had a few limitations. First, the cross-sectional design inherently limits the possibility of causal inferences. Second, the use of a self-reported questionnaire to estimate the number of falls in the previous year is subject to participants’ recall bias. Third, though missing data was handled appropriately, there was a large number of key variables missing from some of the participants. Finally, the results of this study have limited generalizability to other CKD patients who are not currently undergoing HD.

The findings of this study have important implications for the assessment and management of ESRD patients. The results suggest that clinicians should regularly assess patients’ muscle strength and function to determine sarcopenia status and patients’ recent history of falls should be documented, especially those who are 60 years and older and who have a history of cerebrovascular disease. Future studies are necessary to assess the longitudinal impact that low muscle strength and function have on falls risk in this population.

### Conclusion

We conclude that there was no association found between muscle strength and mass parameters with a history of falls in ESRD patients with HD. Patients who reported falls in the previous year were found to be older and to have a history of stroke. Future studies are needed in order to elucidate the factors associated with falls risk in these patients.

## Data Availability

All data produced in the present study are available upon reasonable request to the authors.

## Acknowledgments

The authors would like to thank the members of the GEPENSE study group for their valuable support in data collection. We would also like to thank Hospital Santa Casa de Maringá for their support and for providing access to their facilities. We also acknowledge the Conselho Nacional de Desenvolvimento Científico e Tecnológico (CNPq) and Coordenação de Aperfeiçoamento de Pessoal de Nível Superior (CAPES) for their financial support.

## Notes

**Funding agencies:** Conselho Nacional de Desenvolvimento Científico e Tecnológico (CNPq); Coordenação de Aperfeiçoamento de Pessoal de Nível Superior (CAPES). This manuscript is based on the work presented at the X Congresso Brasileiro de Metabolismo, Nutrição e Exercício (CONBRAMENE); Londrina, Paraná, Brasil; May 15^th^, 2025.

### Competing Interest Statement

The authors have declared no competing interest.

### Funding Statement

This study was funded by the Conselho Nacional de Desenvolvimento Cientifico e Tecnologico (CNPq) and the Coordenacao de Aperfeicoamento de Pessoal de Nivel Superior (CAPES).

### Author Declarations

Comite Permanente de Etica em Pesquisa com Seres Humanos of Universidade Estadual de Maringa gave ethical approval for this work.

